# Multimodal Pain Recognition in Postoperative Patients: A Machine Learning Approach

**DOI:** 10.1101/2023.06.07.23291094

**Authors:** Ajan Subramanian, Rui Cao, Emad Kasaeyan Naeni, Syed Amir Hossein Aqajari, Thomas D. Hughes, Michael-David Calderon, Kai Zheng, Nikil Dutt, Pasi Liljeberg, Sanna Salanterä, Ariana M. Nelson, Amir M. Rahmani

## Abstract

**Objective:** To develop and evaluate a multimodal machine learning-based objective pain assessment algorithm on data collected from post-operative patients.

**Methods:** The proposed method addresses the major challenges that come with using data from such patients like the imbalanced distribution of pain classes and the scarcity of ground-truth labels. Specifically, we extracted automatic features using a convolutional autoencoder (AE) along with data augmentation techniques like weak supervision and minority oversampling to improve our models’ predictive performance. This method was used in conjunction with four different machine learning classifiers: Adaptive Boosting (AdaBoost), Random Forest (RF), Support Vector Machine (SVM), and K-Nearest Neighbors (KNN) to perform binary classification on three increasing levels of pain when compared to no pain.

**Results:** Our models are able to recognize different pain levels with an average balanced accuracy of over 80%.

**Conclusion:** This is the first multimodal pain recognition work done on postoperative patients and our proposed method provides valuable insights for automatic acute pain recognition in such patients.

## Introduction

Pain is defined by the International Association for the Study of Pain (IASP) as “an unpleasant sensory and emotional experience associated with actual or potential tissue damage or described in terms of such damage” [1]. Pain is a unique phenomenon that individuals experience and perceive independently. Younger et al. [2] stated that pain is a subjective experience for which there is no current objective measure. Pain may be classified as either acute or chronic; Kent et al. [3] described acute pain as encompassing the immediate, time-limited bodily response to a noxious stimulus that triggers actions to avoid or mitigate ongoing injury. Chronic pain was first defined loosely by Bonica [4] as pain that extends beyond an expected timeframe; currently, chronic pain is defined as “persistent or recurrent pain lasting longer than three months” [5]. The focus of this article is on acute pain.

Acute pain is a common experience in the post-anesthesia care unit (PACU) in the immediate period following surgery. According to Chou et al. [6], pain occurs in 80% of patients following surgery, and 75% of patients with pain report their pain as either moderate, severe, or extreme. Current guidelines for the assessment of pain in the PACU recommend using a Numerical Rating Scale (NRS) or Verbal Rating Scale (VRS) for patients who are sufficiently awake and coherent to reliably report pain scores [7]. However, Herr et al. [8] identified several patient populations who are at risk for being incapable of providing self-report scores of pain; specifically, these populations include the pediatric population who have yet to develop adequate cognition, elderly patients with dementia, individuals with intellectual disabilities, and those who are unconscious, critically ill, or terminally ill. In these patient populations, Small et al. [7] recommend the use of behavioral pain scales, such as the Pain Assessment in Advanced Dementia (PAINAD), Critical Care Pain Observation Tool (CPOT), or Behavioral Pain Scale (BPS). Despite the pain assessment measures of self-report and behavioral pain scales, each of these methods may be prone to biases. For example, Craig et al. [9] discussed how self-report may be a means to obtain a particular goal that can be influenced by the individual reporting pain. Additionally, Hadjistavropoulos et al. [10] provided the Communications Model of Pain which provided a basis for how expressive behaviors are decoded by observers of individuals in pain, which are influenced by the message clarity transmitted by the individual in pain as well as the unique biases (e.g., knowledge level, assessment skills, and predisposing beliefs) of the individual assessing pain. The difficult nature of interpreting pain scores has resulted in disparities in pain management in minority populations, with research by Staton et al. [11] showing that the black race is a significant predictor of the underestimation of pain by physicians.

Multimodal pain assessment represents one potential method of circumventing the limitations of traditional self-report and behavioral pain assessment tools and an opportunity for enhancing pain assessment in vulnerable populations. Instead of having to rely on only one dimension of pain assessment, such as behaviors through the use of the CPOT or BPS scales, future multimodal pain assessment will incorporate physiological indicators, such as electrodermal activity (EDA), electrocardiogram (ECG), electroencephalogram (EEG) and electromyogram (EMG) as well as behaviors (e.g. facial expression), and perhaps other as-yet undiscovered parameters to capture pain assessment in patient populations that might not be best represented by current assessment strategies. For example, a study by Gélinas et al. [12] found that revisions to the CPOT were necessary because some brain-injured patients may not exhibit certain behaviors that are contained in the CPOT. Similarly, for individuals diagnosed with dementia, Achterberg et al. [13] stated that there is a preponderance of observer-based pain assessment tools, however, these tools retain significant differences between them, as well as concerns for lack of reliability, validity, and sensitivity of change. Enhancing pain assessment through the combination of traditional pain assessment methods with novel multimodal approaches may serve to eventually enhance pain assessment in a greater majority of vulnerable patient populations.

With the advent of connected Internet-of-Things (IoT) devices and wearable sensor technology, automated data collection may achieve continuous pain intensity measurement. A significant amount of research has been conducted in recent years which has sought to develop methods of continuous, automatic, and multimodal pain assessment. For example, prior work conducted by Walter et al. [14] and Werner et al. [15] used skin conductance level (SCL), ECG, electroencephalogram (EEG), and EMG to monitor pain in response to thermal pain. Other works, such as Hammal et al. [16] and Werner et al. [17] have incorporated facial expression monitoring as an indicator of pain. While these studies were immensely beneficial to the scientific community in terms of their contributions to a better understanding of techniques to obtain continuous pain assessment, the setting of these experiments was in highly controlled laboratory environments from healthy participants. Collecting data in real-world situations as opposed to a laboratory setting would allow the researchers to assess a pain assessment technique’s potential in relation to actual pain brought about through a surgical procedure instead of induced pain.

To the best of our knowledge, this is the first work proposing a multimodal pain assessment framework for post-operative patients. It should be noted that a pain assessment study on real patients is associated with several challenges (e.g., imbalanced label distribution, missing data, motion artifacts, etc.) since several parameters such as the intensity, distribution, frequency, and time of the pain as well as the environment cannot be controlled by researchers. Our main contributions are four-fold:

- We conducted a clinical study for multimodal signal acquisition from an acute pain unit of the University of California, Irvine Medical Center (UCIMC)
- We propose a multimodal pain assessment framework using our database (iHurt Pain DB) collected from postoperative patients while obtaining a higher accuracy compared to existing works on healthy subjects [17].
- We use both handcrafted (HC) and automatically generated features outputted from deep learning networks to build our models.
- We provide a novel method to mitigate the presence of sparse and imbalanced labels (due to the real clinical setting of the study) using weak supervision and minority oversampling.

The subsequent sections are organized as follows. Section three talks about related research on this topic. Section four gives a brief overview of our study design. Sections five, six, and seven explain our multimodal framework, the experiments we performed, and the discussion of their results. Finally, Section Eight concludes the article and outlines future research directions.

### Background and Related Work

Current pain assessment (PA) methods rely on caregivers asking patients to self-report their pain levels or observing behavioral or physiological pain responses and using context from the causes of pain. This assessment is often subjective in nature and is affected by social and personal factors including anxiety, depression, disability, and medication. Therefore, there is a pressing need to build an objective pain monitoring system that can predict pain intensity based on physiological factors [18].

The first step in designing such a system is to objectively measure behavioral and/or physiological responses to pain. Behavioral responses are used as protective mechanisms to bring attention to the source of pain and are communicated through facial expressions, body movements, and vocalizations [19]. The use of facial expressions for pain assessment has been studied in depth and is typically examined using the Facial Action Coding System (FACS) [20], which breaks down expressions as movements of elementary Action Units (AUs) based on muscle activity. Facial expressions in response to pain are often varied and may co-occur with other emotions due to the subjective nature of pain experienced in a patient [21]. Such responses can be measured using EMG. Physiological responses to pain stimuli are reflected in the autonomic nervous system’s activities and can be measured through signals like ECG, EDA, and respiratory rate (RR) [22, 23, 24, 25, 26].

To build objective pain monitoring systems it is also very important to consider the type of subjects being recruited because the intensity of pain experienced is highly varied across different groups of people. Prior studies have focused on inducing pain in healthy subjects to reduce the impact of pre-existing conditions that might inject biases into the data (Biovid, BP4D, MIntPAIN, SenseEmotion, X-ITE Pain) [14, 27, 28, 29, 30], whereas some studies have focused on patients with chronic pain (EmoPain, and UNBC McMaster database) [19, 31]. In clinical settings, many patients suffer from ongoing chronic pain without the involvement of external stimuli, but pain response is oftentimes intensified through necessary medical procedures like surgeries. An underrepresented population in pain studies is patients suffering from acute post-operative pain. Our prior work has focused on building pain assessment models on single modalities like ECG [29], EDA [33], and PPG [34] from our postoperative pain study. Even though the results we achieve for each of these single modalities are significant, we still do not leverage the multimodal nature of our collected dataset. Building models using a single modality might not be able to capture the full extent of a patient’s painful experience and often has caveats in some clinical contexts. Heterogeneous sources of data, on the contrary, could complement each other and lead to improved performance over any single modality. Therefore, building a multimodal pain assessment system that utilizes both physiological and behavioral responses to pain can prove to be vital for vulnerable patient populations.

## Methods

### iHurt Study design

We conducted a biomedical data collection study on 25 post-operative patients reporting various degrees of pain symptoms. Multimodal biosignals (ECG, EMG, EDA, PPG) were collected from patients likely having mild to moderate pain, who were asked to perform a few light physical activities while acquiring data. We also collected primary demographic information from each patient including height, weight, sex, and body mass index. All signals were collected using the iHurt system.

### iHurt system

iHurt is a system that measures facial muscle activity (i.e., changes in facial expression) in conjunction with physiological signals such as heart rate, heart rate variability, respiratory rate, and electrodermal activity for the purpose of developing an algorithm for pain assessment in hospitalized patients. The system uses the two following components to capture raw signals.

1. Eight-Channel Biopotential Acquisition Device: Our team at the University of Turku, Finland developed a biopotential acquisition device to measure ECG and EMG signals. The device incorporates commercially available electrodes, electrode-to-device lead wires, an ADS1299-based portable device, and computer software (LabVIEW version 14.02f, National Instruments) to visualize data streaming from the portable device. Raw signals from the electrodes are sampled at 500 samples per second and are sent to the computer software via Bluetooth for visualization [35].
2. Empatica E4: We use the commercially available Empatica E4 wristband (Empatica Inc, Boston, MA, USA) [33] to measure EDA and PPG signals. The purpose of using a wristband was to allow our participants to move freely without any impediments. The Empatica E4 was connected to the participants’ phones over Bluetooth for visualization.

This is the first claimed study that collected biosignals from postoperative adult patients in hospitals. All participants (age: 23 - 89 years) were recruited from the University of California, Irvine Medical Center after obtaining Institutional Review Board approval (IRB, HS: 2017-3747). The patients were recruited to the study from July 2018 to October 2019. We removed 3 participants’ data from the final dataset due to the presence of excessive motion artifacts. We also excluded 2 additional patients since they were wearing the Empatica E4 watch on their arm that received IV (intravenous) medication. This resulted in unreliable EDA signals due to conditions like skin rash and itching. This left us with data from 20 patients to build our pain recognition system. The dataset also contains rich annotation with self-reported pain scores based on the 11-point Numeric Rating Scale (NRS) from 0 – 10. A detailed explanation of the dataset and the study design can be found in [37]. We intend to make the de-identified dataset available to the research community for further analysis and applications.

### Data Processing Pipeline

The first step in building our multimodal pain assessment system was to process the raw signals collected during trials. The data processing pipeline consisted of the following steps:

- We filtered the signal to remove powerline interference, baseline wander, and motion artifact noise.
- We performed feature extraction on the filtered signals to obtain amplitude and variability features in the time domain. The time domain features were extracted using 5.5-second and 10-second windows. The 5.5-second window size was extracted to compare with prior work [17].
- In addition to HC features, we also used automatic features which were outputted from a deep neural network.
- Once the features were extracted, we tagged them with their corresponding labels based on the nearest timestamp of the label.

Each of these processing steps was applied individually to each of the four modalities. Processed data from each of the modalities were combined using either early fusion (EF) or late fusion (LF). The types of HC features extracted from each modality and the deep learning pipeline for extracting automatic features are described in detail.

### ECG Handcrafted Features

The ECG channel was filtered using a Butterworth band-pass filter with the frequency ranges of [0.1,250] Hz. The HRV HC features were extracted with pyHRV, an open-source Python toolbox [38] using the R-peaks extracted from the ECG signal via a bidirectional long short-term memory network [39]. These features were extracted from two window sizes, 5.5 and 10 seconds. There were 19 time-domain (TD) features. The TD features extracted from NN intervals, or the time interval between successive R-peaks, comprised of the slope of these intervals, 5 statistical features (total count, mean, minimum, maximum, and standard deviation), 9 difference features (mean difference, minimum difference, maximum difference, standard deviation of successive interval differences, root mean square of successive interval differences, number of interval differences greater than 20ms and 50ms, and percentage of successive interval differences that differ by more than 20ms and 50ms), and 4 heart rate features (mean, minimum, maximum, and standard deviation).

### EMG Handcrafted Features

The preprocessing phase of EMG channels comprised of a 20Hz high pass filter and two notch filters at 50Hz and 100Hz all using a Butterworth filter. Like ECG features, we extracted EMG features from 5.5 and 10-second windows on 5 different channels for each major facial muscle. The 10 amplitude features extracted were 1) peak, 2) peak-to-peak mean value (p2pmv), 3) root mean squared (rms), 4) mean of the absolute values of the second differences (mavsd), 5) mean of the absolute values of the first differences (mavfd), 6) mean of the absolute values of the second differences of the normalized signal (mavsdn), 7) mean of the absolute values of the first differences of the normalized signal (mavfdn), 8) mean of local minima values (mlocminv), 9) mean of local maxima values (mlocmaxv), and 10) mean of absolute values (mav). The 4 variability features were 1) variance, 2) standard deviation, 3) range, and 4) interquartile range. All 14 features were calculated for 5 different EMG channels resulting in 70 EMG features in total.

### EDA Handcrafted Features

We used the pyEDA library [40] for pre-processing and feature extraction of EDA signals. In the pre-processing part, first, we used a moving average across a 1-second window to remove the motion artifacts and smooth the data. Second, a low-pass Butterworth filter on the phasic data was applied to remove the line noise. Lastly, preprocessed EDA signals corresponding to each different pain level were visualized to ensure the validity of the signals. In the feature extraction part, the cvxEDA algorithm [41] was employed to extract the phasic component of EDA signals. The EDA signals’ peaks or bursts are considered variations in the phasic component of the signal. Therefore, the clean signals and extracted phasic component of signals were fed to the statistical feature extraction module to extract the number of peaks, the average value, and the maximum and minimum value of the signals. Moreover, these extracted features were further employed in the post-feature extraction module to extract 8 more features: (1) the difference between the maximum and the minimum value of the signal, (2) the standard deviation, (3) the difference between the upper and lower quartiles (4) root mean square, (5) the mean value of local minima, (6) the mean value of local maxima, (7) the mean of the absolute values of the first differences, and (8) the mean of the absolute values of the second differences. This resulted in 12 EDA features in total.

### PPG-based Respiratory Rate Handcrafted Features

We pre-processed the PPG signal before extracting the respiratory rate from it. Two filters were used during the preprocessing. We first used a Butterworth bandpass filter to remove noises including motion artifacts. Then, a moving average filter was implemented to smooth the PPG signal. After that, we applied an Empirical Mode Decomposition (EMD) based method proposed by Madhav et al. [42] to derive respiration signals from filtered PPG signals. This method was proven to derive RR from a PPG signal with high accuracy (99.87%). Ten features were extracted from the respiratory signal including (1) the number of inhale peaks, (2) the mean value of the signal, (3) the maximum value (4) the minimum value (5) the difference between the maximum and the minimum value, (6) standard deviation, (7) the average value of the inhale peak intervals, (8) the standard deviation of the inhale peak intervals, (9) the root mean square of successive differences between adjacent inhale peak intervals, (10) standard deviation of inhale duration/average inhale duration. A visualization of the HC feature pipeline is shown in Fig 1.

**Fig 1.**
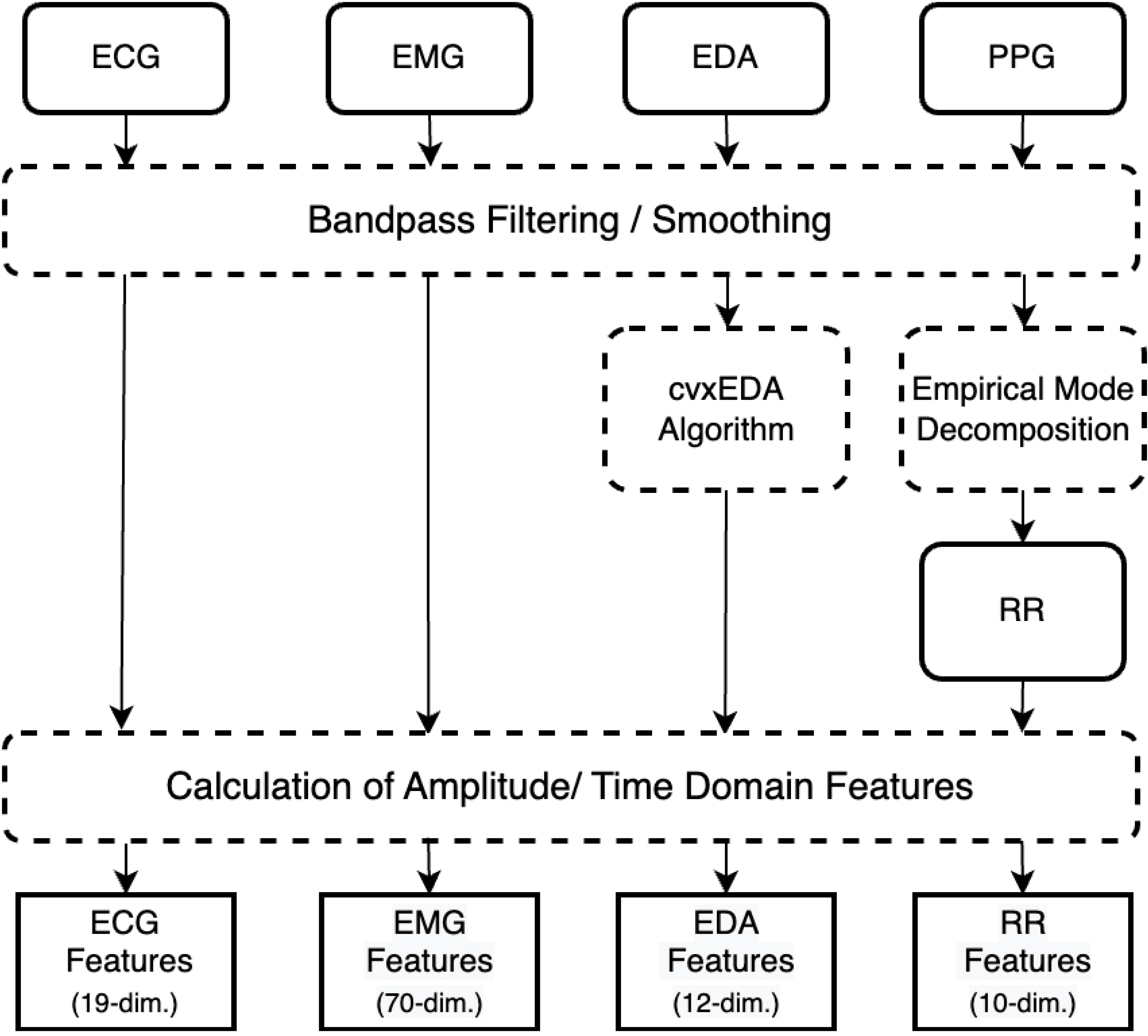
Handcrafted feature extraction pipeline.

### Automatic Feature Extraction Pipeline

As the dimensionality of biomedical data increases, it becomes increasingly difficult to train a machine learning algorithm on the entire uncompressed dataset. This often leads to a large training time and is computationally more expensive overall. One possible solution is to perform feature engineering to get a compressed and interpretable representation of the signal. Another alternative approach, however, is to use the compressed or latent representation of that data obtained from deep learning networks trained for that specific task. Using automatic features helps in dimensionality reduction and can provide us with a sophisticated yet succinct representation of the data that HC features alone cannot provide. This automatic feature extraction is typically carried out by an autoencoder (AE) network, which is an unsupervised neural network that learns how to efficiently compress and encode the data into a lower-dimensional space [43, 44]. Autoencoders are composed of two separate networks, an encoder, and a decoder. The encoder network acts as a bottleneck layer and maps the input into a lower-dimensional feature space. The decoder network tries to reconstruct this lower-dimensional feature vector into the original input size. The entire network is trained to minimize the reconstruction loss (i.e., mean-squared error) by iteratively updating its weights and biases through backpropagation.

A convolutional AE from the pyEDA library was used to extract automatic features. Fig 2 shows the architecture of the AE. First, a linear layer (L1) is used to downsample the input signal with Input Shape length to a length that is the closest power of 2 (CP2). This was done to make the model scalable to an arbitrary input size. The encoder half of the network consists of three 1-D convolutional layers (C1, C2, and C3) and a linear layer (L2) which flatten and downsamples the input vector to a lower-dimensional latent vector. The number of dimensions of this latent vector (Feature Size) corresponds to the number of automatic features extracted and was set prior to training the network. A total of 32 features were extracted from ECG, EDA, and RR signals. Whereas a total of 30 features were extracted from the EMG signal (6 features from each of the 5 channels). The decoder half of the network consists of three 1-D de-convolutional layers (DeC1, DeC2, and DeC3) to reconstruct the input signal from the latent vector. A final linear layer (L3) is then used to flatten and reconstruct the signal to its original dimension. Both encoder and decoder networks have ReLU (Rectified Linear Unit) activation between layers. Window sizes of both 5.5 and 10 seconds were applied to the filtered signals. This was done to compare the performance with HC features. After signals from each of the modalities were normalized, they were trained on separate AE models for each modality. In addition to the convolutional AE, we also extracted features from an LSTM (long-short-term memory) AE network. This resulted in two different feature extraction methods (convolutional and LSTM) that spanned two different window lengths (5.5 and 10 seconds).

**Fig 2.**
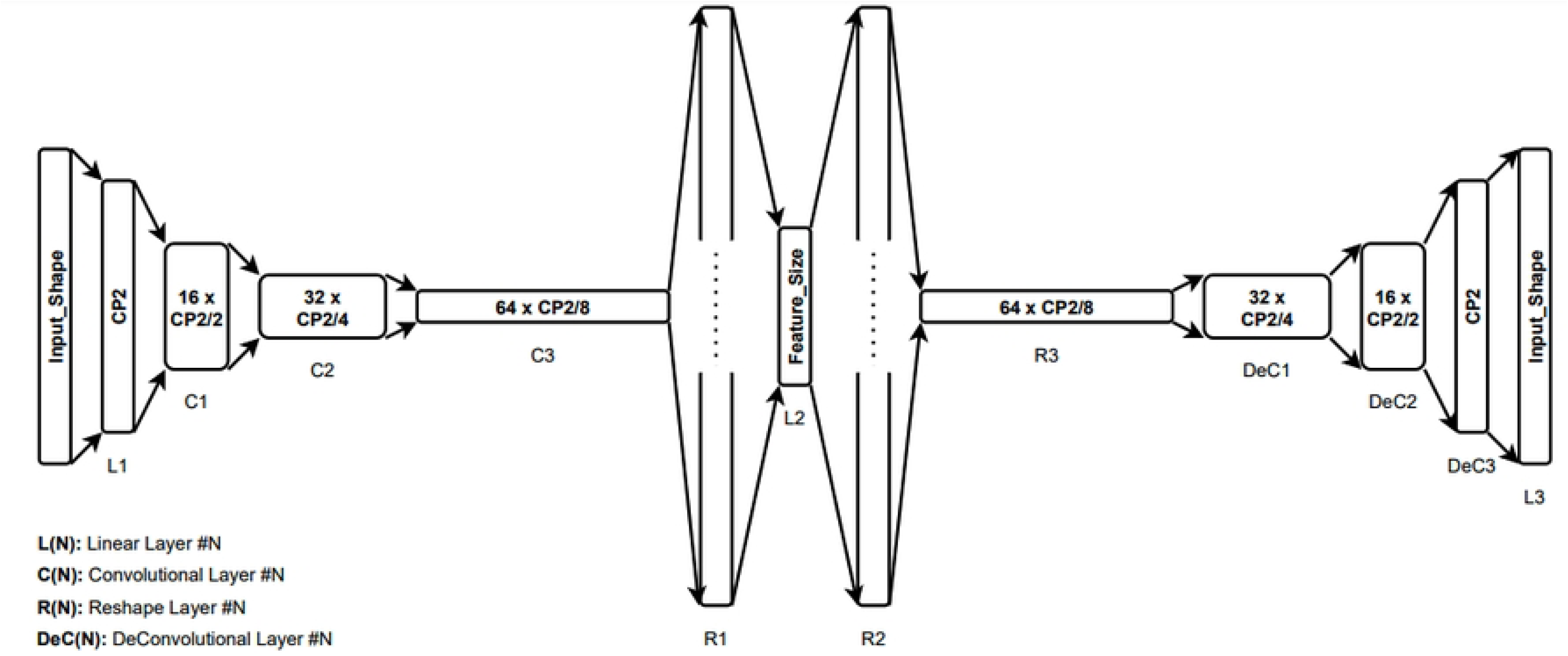
The architecture of the pyEDA convolutional autoencoder.

The batch size was set to 10, the number of training epochs was set to 100, and the ADAM optimizer [42] was used with a learning rate of 1e-3. A total of 126 feature vectors across all 4 modalities were extracted from each AE network. A visualization of our automatic feature extraction pipeline is shown in Fig 3.

**Fig 3.**
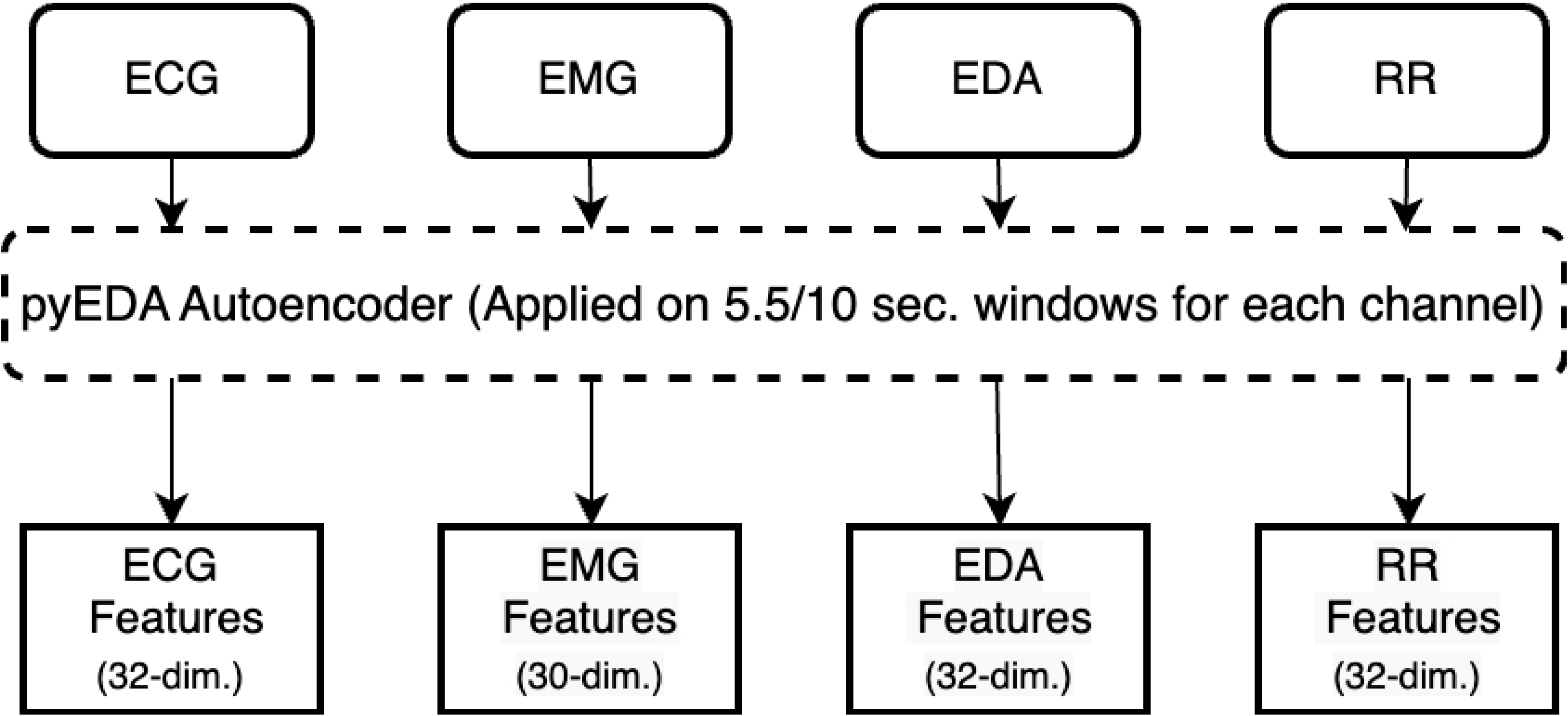
Automatic feature extraction pipeline.

### Data Augmentation

There were several inherent challenges in the distribution of labels as NRS values recorded during the clinical trials of this study were collected from real postoperative patients. This problem bears less significance while studying healthy participants since the stimulated pain can be controlled during the experiments. Consequently, occurrences of some pain levels far exceeded those of others. For example, among all patients, there were only 4 reported occurrences of pain level 10, whereas there were more than 80 reported occurrences of pain level 4. This imbalanced distribution was inevitable due to the subjective nature and the different sources of pain among the participants. Therefore, while downsampling our pain labels to 4 classes, thresholds for each downsampled class were carefully chosen to ensure a more evenly distributed set of labels. The pain levels ranged from a baseline level of pain (BL) or no pain to 3 increasing intensities of pain (PL 1-3). The thresholds for the pain levels were as follows −1) PL1 ranged from 0 to 3, 2) PL2 ranged from 4 to 6, and 3) PL3 ranged from 7 to 10. All the ranges here are inclusive.

Since we asked patients to report their pain levels only while they performed pain-inducing activities, the number of labels generated was sparse. Both HC and automatic features were combined with the corresponding labels using timestamps that were within the nearest 5.5 or 10 seconds (labeling threshold) of the reported NRS value. This depended on the window size of the features extracted. Because of having sparse labels, many of the feature windows were not assigned a corresponding label. To mitigate the problem of having an imbalanced and sparse label distribution, two techniques were exploited.

1. Minority Oversampling: The first technique, called Synthetic Minority Oversampling (Smote), is a type of data augmentation that over-samples the minority class [46]. Smote works by first choosing a minority class instance at random and finding its k nearest minority class neighbors. It then creates a synthetic example at a randomly selected point between two instances of the minority class in that feature space. The experiments involving Smote were implemented using the imbalanced-learn Python library [47].
2. Weak Supervision: The second technique we utilized is weak supervision using the Snorkel framework [48]. Rather than employing an expert to manually label the unlabeled instances, Snorkel allows its users to write labeling functions that can make use of heuristics, patterns, external knowledge bases, and third-party machine learning models. Weak supervision is typically employed to label large volumes of unlabeled data when there are noisy, limited, or imprecise sources. For our pain assessment algorithm, we decided to use third-party machine learning models to label the remaining unlabeled instances. All the data points that were within the labeling threshold were considered as “strong labels”, or ground-truth values collected from patients during trials. The remaining unlabeled data points were kept aside for Snorkel to provide a weakly supervised label. The strong labels were fed into Snorkel’s labeling function consisting of three off-the-shelf machine learning models: (i) a Support-Vector Machine (SVM) with a radial basis function kernel, (ii) a Random Forest (RF) classifier, and (iii) a K-Nearest Neighbor (KNN) classifier with uniform weights. Once each model was trained on the strong labels, it was used to make predictions on the remaining unlabeled data. The predictions from these three models were collected and converted into a single confidence-weighted label per data point using Snorkel’s “LabelModel” function. This function outputs the most confident prediction as the label for each data point. To perform a fair assessment of the reliability and accuracy of our algorithm, we used Smote and Snorkel only while training our machine learning models. The performance of these models was measured solely on ground-truth (strong) labels collected during trials. This way, there is no implicit bias introduced from mislabeling or upsampling certain data points to skew model predictions.

### Multimodal Machine Learning Models

To compare the performance of our multimodal machine learning models with the prior work, we performed binary classification using a leave-one-subject-out cross-validation approach [49]. In this method, a model’s performance is validated over multiple folds in such a way that data from each patient is either in the training set or in the testing set. The purpose of using this method is to provide generalizability to unseen patients and to avoid overfitting by averaging the results over multiple folds. The eventual goal of this study is to build personalized models that make predictions on a single patient but learn from data collected from a larger population of similar patients. The following machine learning models were used to evaluate the performance of our pain assessment algorithm: (1) K-nearest neighbors, (2) Random Forest classifier, (3) AdaBoost (Adaptive Boosting), (4) and an SVM (Support Vector Machine). The models were then evaluated using leave-one subject-out cross-validation. Four separate models were trained for each of the three pain intensities (e.g., BL, no pain versus PL1, the lowest pain level, or BL vs PL3, the highest pain level).

### Fusing Modalities

Two fusion approaches were used while combining features across different modalities. The first one is early or feature-level fusion which concatenates feature vectors across different modalities based on their timestamps. The resulting data that is now higher in dimension than any one single modality is then fed into our classifier to make predictions. While concatenating features across different modalities, a threshold of either 5.5 or 10 seconds was used to combine the modalities depending on the features extracted. The second approach was late or decision-level fusion where each modality is fed to a separate classifier and the final classification result is based on the fusion of outputs from the different modalities [50].

### Feature Selection

Since there were a lot of features generated during the data processing phase, we had to select a subset of the most informative features to build our models with. Therefore, to reduce the complexity and training time of the resulting model, feature selection using Gini importance was performed. Gini importance is a lightweight method that is simple and fast to compute. Since we extracted a relatively large number of features in our method, it made sense to use a computationally low-cost algorithm for feature selection. We computed the Gini importance of the features from the data in the training fold with the help of a random forest classifier and selected the top 25 features. We then trained our model on these top 25 features and evaluated them in the validation fold. Our proposed multimodal pain recognition system is shown in Fig 4.

**Fig 4.**
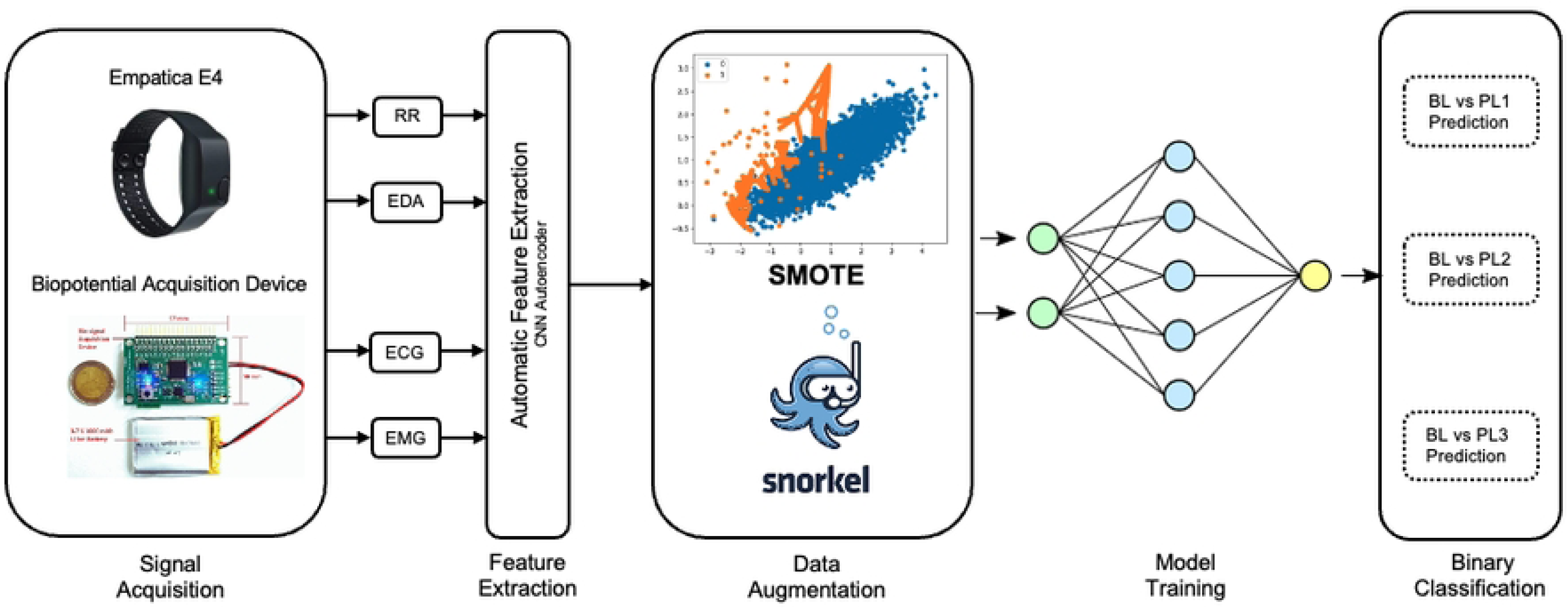
Proposed multimodal pain recognition system.

## Results

### Experimental Settings

The goal of our experiments was to compare the performance of using only a single modality to build our models over using a combination of multiple modalities. We trained several different models for each of the pain intensities that varied in the types of modalities, data augmentation techniques, machine learning models, and fusion techniques used. Fig 5 shows the general pipeline of the experiments we conducted. We first select the type of modalities to train on, which varied from only using each of the single modalities separately to using a combination of all 4 modalities. Moreover, these modalities varied on the types of features used, like HC or automatic features. In the case of using multiple modalities, we had two choices of fusion: early (Fig 5 left) and late (Fig 5 right). These architectures varied in how the modalities were combined, either before training (early), or at the decision level (late) after training using majority voting. The data preparation process involved feature selection and data augmentation. These models could either be trained with no data augmentation, with just Smote or Snorkel, or a combination of both. The last step of the pipeline before making predictions involved choosing the type of machine learning algorithms, like SVM, Random Forest (RF), Adaptive Boosting (AdaBoost), or K-Nearest Neighbors (KNN). Due to the lack of space, only the best-performing single and multimodal model configurations are mentioned in the section below.

**Fig 5.**
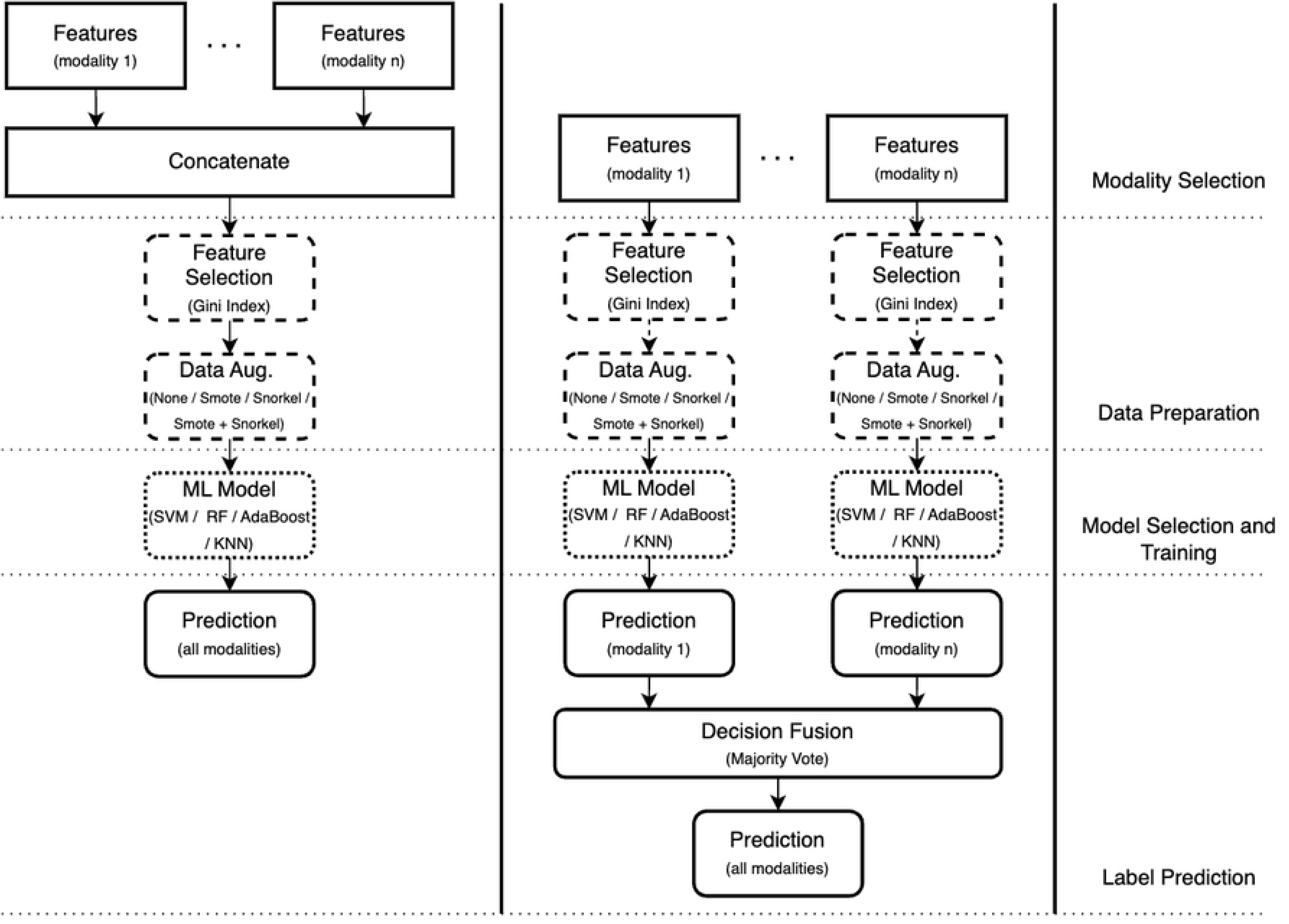
Our proposed general multimodal pipeline based on early fusion (left) and late fusion (right).

### Experimental Results

Tables 1 and 2 present the best-performing single-modal and multimodal models for each of the three pain intensities. For comparison, the best multimodal results from Werner et al. [17], Martinez et al. [24], Wang et al. [25], and Subramaniam et al. [26] are also mentioned. We use balanced accuracy as an evaluation criterion because our dataset had an imbalanced class distribution. Balanced accuracy is defined as the average of the true positive rate and the true negative rate.

**Table 1.** Best scores: single modality and multiple modalities.

| Pain Levels | ECG Scores | EMG Scores | EDA Scores | RR Scores | Multiple Modality |
| --- | --- | --- | --- | --- | --- |
| BL vs. PL1 | 82.14 | 86.0 | 79.18 | 84.62 | 82.14 |
| BL vs. PL2 | 86.11 | 84.53 | 82.94 | 88.24 | 86.11 |
| BL vs. PL3 | 75.0 | 78.12 | 75.0 | 76.23 | 75.0 |
| Mean | 81.08 | 82.88 | 79.04 | 83.03 | 81.08 |
| Classifier Config. | LSTM AE (10s), Strong, SVM | HC (10s), Snorkel, SVM | CNN AE (10s), Strong, SVM | HC (10s), Strong, SVM | EF, LSTM AE (10s), Strong, SVM |

**Table 2.** Multiple modalities: comparison with other methods.

|  | Werner et al. [17] | Martinez et al. [24] | Wang et al. [25] | Subramaniam et al. [26] | Our Method |
| --- | --- | --- | --- | --- | --- |
| Mean Scores | 65.27 | 66.84 | 70.28 | 76.28 | 81.08 |
| Modalities | Video, ECG, EMG, EDA | ECG, EMG, EDA | ECG, EMG, EDA | ECG, EDA | ECG, EMG, EDA, RR |

## Discussion

### Principal Results

From the single modality results (Table I), it is evident that RR models outperform all other modalities, especially for the BL vs PL1 and BL vs PL2 models. Overall, models from all modalities have relatively lower scores in the BL vs PL3 category. The comparatively lower performance of EDA models over other modalities suggests that variations in EDA signal response to different pain levels are more difficult to distinguish. From our experiments, the best-performing multimodal model was trained on automatic features outputted from our LSTM network with 10-second window size. This model made use of strong labels without any data augmentation techniques. It should be noted that the best-performing ECG and multimodal models have the same results and share identical configurations. It is very likely that the ECG features influenced the performance of the multimodal model.

The relatively poor performances of the BL vs PL1 and BL vs PL3 models across both single and multimodal models are also understandable because they lie at the extremes of the pain threshold. The BL vs PL1 models might find it more challenging to distinguish between baseline levels and the lowest pain intensity due to the subtlety of the physiological responses collected while experiencing this pain level. The BL vs PL3, however, might find it challenging to distinguish pain levels due to the scarcity of such labels collected during trials. Data augmentation can help mitigate this problem, but there is no substitute for real data. On the contrary, the BL vs PL2 models performed better due to the relative abundance of such labels reported during trials.

In terms of modalities, the best-performing model uses RR alone. However, for the last pain category, the EMG model outperformed all the other models. One justification for these results could be due to the dynamic nature of these signals in response to pain stimuli. Since we were able to effectively isolate and capture periods of higher pain intensity with smaller window sizes, this could help the models better distinguish between baseline and other pain levels.

The best-performing multimodal models use early fusion or feature-level fusion. One intuition as to why early fusion might perform better overall is due to the detection of correlated features across modalities obtained after using feature selection [51]. Late fusion, on the contrary, builds independent models for each modality and fuses them based on their predictions using majority voting. Therefore, by treating each modality as independent, there is a potential loss of correlation in the combined feature space.

Overall, the single modality results, specifically RR, outperform the multimodal models in all categories. This has not been the case in prior studies done on healthy subjects. But our experiments suggest that a combination of multiple modalities in data collected from postoperative patients has the potential to skew results. Since there is a risk of missing data and noise in our signals, it is imperative to carefully align them when combining the modalities. Multiple modalities certainly have the potential to add more useful information over a single modality and can be used to introduce complementary information and resiliency when any one modality fails or is too noisy [52]. They are also more robust and can improve generalizability when patients experience different types of pain and in varying degrees. However, there are also advantages to using a single modality. They are simpler and easier to interpret when measuring each feature’s contribution to the output. This also reduces computational complexity and training time. While comparing our results to [17, 24, 25, 26], it can be observed that our models outperform their models in mean pain assessment scores. However, this is not entirely a fair comparison because we use 3 pain levels instead of 4 and our patients are not healthy.

One of the main research directions we would like to explore in the future is to build real-time multimodal pain assessment systems using deep learning architectures. In such scenarios, it is quite possible to have missing or incomplete data from or more modalities. Moreover, real-time systems are limited by their computational complexity and power constraints. Therefore, with the help of the experiments performed in this study, we hope to build models that can dynamically determine which modalities to use in an energy-efficient manner without compromising performance given the clinical context.

### Limitations

The main limitation of our algorithm is the presence of noise in the form of motion artifacts produced while collecting physiological signals. Since we obtained data from real postoperative patients in a clinical setting, they were allowed to move more freely in comparison to experiments performed in laboratory settings. The presence of these motion artifacts diminished the quality of our data, and thus negatively impacted our machine-learning algorithms. We also must acknowledge the more complicated facets of pain that are not fully captured by our algorithm like the number of days post-surgery, the amount of pain medication dosing, the location, and the type of pain experienced. We would like to account for these factors while conducting future studies.

## Conclusion

In this article, we presented a multimodal machine-learning framework for classifying pain in real post-operative patients from the iHurt Pain Database. Both traditional handcrafted features and deep learning-generated automatic features were extracted from physiological signals (ECG, EDA, EMG, PPG). We conducted several experiments to perform binary classification among three different pain intensities vs baseline levels of pain. Models for each of these intensities were varied based on the modalities used, the different types of data augmentation techniques (Smote, Snorkel, or both), the machine learning algorithms used, and the type of modality fusion used. Our results showed that binary pain classification greatly benefits from using data augmentation techniques in conjunction with automatic features. The single-modality models from RR and EMG outperformed the multimodal models. The BL vs PL3 model with the best results was trained on EMG data alone, which suggests that facial muscle activation can play a vital role in distinguishing higher pain intensities from baseline levels of pain. This is consistent from a clinical perspective because higher pain intensities are more commonly associated with acute pain.

However, since pain is a subjective experience that tends to have a large inter-individual variability, building a monolithic model for all patients might not be a viable solution. A promising future direction for this research study is to build personalized machine learning models that can benefit from using data from groups of similar patients, but which are finetuned to make predictions on a single person. Prior research has used multitask machine learning (MTL) to account for inter-individual variability and build personalized models for the task of mood prediction [53]. This is a feasible future research direction that would be applicable to the domain of pain assessment, not only for the acute pain of surgery but also for patients that experience chronic pain. We believe that personalized modeling will be a vital step in creating clinically viable pain assessment algorithms.

## Data Availability

The data used in this study include sensitive health information that cannot be publicly shared due to ethical restrictions outlined in the informed consent. Permission to use the data was granted by the participants solely for the specific purpose described in the consent. Data access requires individual consent or approval from the Institutional Review Board. Researchers interested in accessing the data should contact the project’s PI, Associate Professor Amir Rahmani, at, located at the University of California, Irvine, Institute for Future Health, Irvine, California, United States, 92617.

## Acknowledgments

N/A

## References

1. Merskey HA. Pain terms: a list with definitions and notes on usage. Recommended by the IASP Subcommittee on Taxonomy. Pain. 1979;6:249–52.

2. Younger J, McCue R, Mackey S. Pain outcomes: a brief review of instruments and techniques. Current pain and headache reports. 2009 Feb;13:39–43. doi:10.1007/s11916-009-0009-x.

3. Kent ML, Tighe PJ, Belfer I, Brennan TJ, Bruehl S, Brummett CM, Buckenmaier III CC, Buvanendran A, Cohen RI, Desjardins P, Edwards D. The ACTTION–APS–AAPM Pain Taxonomy (AAAPT) multidimensional approach to classifying acute pain conditions. Pain Medicine. 2017 May 1;18(5):947–58. doi:10.1093/pm/pnx019.

4. Bonica JJ. Management of cancer pain. In Pain in the Cancer Patient: Pathogenesis, Diagnosis and Therapy 1984 (pp. 13–27). Springer Berlin Heidelberg.

5. Treede RD, Rief W, Barke A, Aziz Q, Bennett MI, Benoliel R, Cohen M, Evers S, Finnerup NB, First MB, Giamberardino MA. A classification of chronic pain for ICD-11. Pain. 2015 Jun;156(6):1003. doi:10.1097/j.pain.0000000000000160.

6. Chou R, Gordon DB, de Leon-Casasola OA, Rosenberg JM, Bickler S, Brennan T, Carter T, Cassidy CL, Chittenden EH, Degenhardt E, Griffith S. Management of Postoperative Pain: a clinical practice guideline from the American pain society, the American Society of Regional Anesthesia and Pain Medicine, and the American Society of Anesthesiologists’ committee on regional anesthesia, executive committee, and administrative council. The journal of pain. 2016 Feb 1;17(2):131–57. doi:10.1016/j.jpain.2015.12.008.

7. Small C, Laycock HJ. Acute postoperative pain management. Journal of British Surgery. 2020 Jan;107(2):e70–80. doi:10.1002/bjs.11477.

8. Herr K, Coyne PJ, Ely E, Gélinas C, Manworren RC. Pain assessment in the patient unable to self-report: clinical practice recommendations in support of the ASPMN 2019 position statement. Pain Management Nursing. 2019 Oct 1;20(5):404–17. doi:10.1016/j.pmn.2019.07.005.

9. Kappesser J. The facial expression of pain in humans considered from a social perspective. Philosophical Transactions of the Royal Society B. 2019 Nov 11;374(1785):20190284. doi:10.1098/rstb.2019.0284.

10. Hadjistavropoulos T, Craig KD. A theoretical framework for understanding self-report and observational measures of pain: a communications model. Behaviour research and therapy. 2002 May 1;40(5):551–70. doi:10.1016/s0005-7967(01)00072-9.

11. Staton LJ, Panda M, Chen I, Genao I, Kurz J, Pasanen M, Mechaber AJ, Menon M, O’Rorke J, Wood J, Rosenberg E. When race matters: disagreement in pain perception between patients and their physicians in primary care. Journal of the National Medical Association. 2007 May;99(5):532.

12. Gélinas C, Boitor M, Puntillo KA, Arbour C, Topolovec-Vranic J, Cusimano MD, Choinière M, Streiner DL. Behaviors indicative of pain in brain-injured adult patients with different levels of consciousness in the intensive care unit. Journal of Pain and Symptom Management. 2019 Apr 1;57(4):761–73. doi:10.1016/j.jpainsymman.2018.12.333.

13. Achterberg W, Lautenbacher S, Husebo B, Erdal A, Herr K. Pain in dementia. Der Schmerz. 2021 Apr;35:130–8. doi:10.1097/PR9.0000000000000803.

14. Walter S, Gruss S, Ehleiter H, Tan J, Traue HC, Werner P, Al-Hamadi A, Crawcour S, Andrade AO, da Silva GM. The biovid heat pain database data for the advancement and systematic validation of an automated pain recognition system. In 2013 IEEE international conference on cybernetics (CYBCO) 2013 Jun 13 (pp. 128–131). IEEE. doi: 10.1109/CYBConf.2013.6617456.

15. Werner P, Al-Hamadi A, Niese R, Walter S, Gruss S, Traue HC. Towards pain monitoring: Facial expression, head pose, a new database, an automatic system and remaining challenges. In Proceedings of the British Machine Vision Conference 2013 Sep (pp. 1–13). doi: 10.5244/C.27.119.

16. Hammal Z, Cohn JF. Automatic detection of pain intensity. In Proceedings of the 14th ACM international conference on Multimodal interaction 2012 Oct 22 (pp. 47–52). doi:10.1145/2388676.2388688.

17. Werner P, Al-Hamadi A, Niese R, Walter S, Gruss S, Traue HC. Automatic pain recognition from video and biomedical signals. In 2014 22nd International Conference on Pattern Recognition 2014 Aug 24 (pp. 4582–4587). IEEE. doi: 10.1109/ICPR.2014.784.

18. Werner P, Lopez-Martinez D, Walter S, Al-Hamadi A, Gruss S, Picard RW. Automatic recognition methods supporting pain assessment: A survey. IEEE Transactions on Affective Computing. 2019 Oct 14;13(1):530–52. doi: 10.1109/TAFFC.2019.2946774.

19 . Aung MS, Kaltwang S, Romera-Paredes B, Martinez B, Singh A, Cella M, Valstar M, Meng H, Kemp A, Shafizadeh M, Elkins AC. The automatic detection of chronic pain-related expression: requirements, challenges and the multimodal EmoPain dataset. IEEE transactions on affective computing. 2015 Jul 30;7(4):435–51. doi:10.1109/TAFFC.2015.2462830.

20. Rosenberg EL, Ekman P, editors. What the face reveals: Basic and applied studies of spontaneous expression using the Facial Action Coding System (FACS). Oxford University Press; 2020 Jun 15. (New York, 2005; online edn, Oxford Academic, 22 Mar. 2012), https://doi.org/10.1093/acprof:oso/9780195179644.001.0001, accessed 24 Feb. 2023.

21. Lopez Martinez D, Picard R. Personalized automatic estimation of self-reported pain intensity from facial expressions. In Proceedings of the IEEE conference on computer vision and pattern recognition workshops 2017 (pp. 70–79).

22. Gruss S, Geiger M, Werner P, Wilhelm O, Traue HC, Al-Hamadi A, Walter S. Multi-modal signals for analyzing pain responses to thermal and electrical stimuli. JoVE (Journal of Visualized Experiments). 2019 Apr 5 (146):e59057. doi:10.3791/59057.

23. Hernandez J, McDuff D, Picard RW. Biowatch: estimation of heart and breathing rates from wrist motions. In 2015 9th International Conference on Pervasive Computing Technologies for Healthcare (PervasiveHealth) 2015 May 20 (pp. 169–176). IEEE. doi: 10.4108/icst.pervasivehealth.2015.259064.

24. Lopez-Martinez D, Picard R. Multi-task neural networks for personalized pain recognition from physiological signals. In 2017 Seventh International Conference on Affective Computing and Intelligent Interaction Workshops and Demos (ACIIW) 2017 Oct 23 (pp. 181–184). IEEE.

25. Wang R, Xu K, Feng H, Chen W. Hybrid RNN-ANN based deep physiological network for pain recognition. In 2020 42nd Annual International Conference of the IEEE Engineering in Medicine & Biology Society (EMBC) 2020 Jul 20 (pp. 5584–5587). IEEE. doi: 10.1109/EMBC44109.2020.9175247.

26. Subramaniam SD, Dass B. Automated nociceptive pain assessment using physiological signals and a hybrid deep learning network. IEEE Sensors Journal. 2020 Sep 11;21(3):3335–43. doi: 10.1109/JSEN.2020.3023656.

27. Zhang X, Yin L, Cohn JF, Canavan S, Reale M, Horowitz A, Liu P, Girard JM. Bp4d-spontaneous: a high-resolution spontaneous 3d dynamic facial expression database. Image and Vision Computing. 2014 Oct 1;32(10):692–706.

28. Haque MA, Bautista RB, Noroozi F, Kulkarni K, Laursen CB, Irani R, Bellantonio M, Escalera S, Anbarjafari G, Nasrollahi K, Andersen OK. Deep multimodal pain recognition: a database and comparison of spatio-temporal visual modalities. In 2018 13th IEEE International Conference on Automatic Face & Gesture Recognition (FG 2018) 2018 May 15 (pp. 250–257). IEEE. doi: 10.1109/FG.2018.00044.

29. Velana M, Gruss S, Layher G, Thiam P, Zhang Y, Schork D, Kessler V, Meudt S, Neumann H, Kim J, Schwenker F. The senseemotion database: A multimodal database for the development and systematic validation of an automatic pain-and emotion-recognition system. In Multimodal Pattern Recognition of Social Signals in Human-Computer-Interaction: 4th IAPR TC 9 Workshop, MPRSS 2016, Cancun, Mexico, December 4, 2016, Revised Selected Papers 4 2017 (pp. 127–139). Springer International Publishing. doi: 10.1007/978-3-319-59259-6_11.

30. Werner P, Al-Hamadi A, Gruss S, Walter S. Twofold-multimodal pain recognition with the X-ITE pain database. In 2019 8th International Conference on Affective Computing and Intelligent Interaction Workshops and Demos (ACIIW) 2019 Sep 3 (pp. 290–296). IEEE. doi: 10.1109/ACIIW.2019.8925061.

31. Lucey P, Cohn JF, Prkachin KM, Solomon PE, Matthews I. Painful data: The UNBC-McMaster shoulder pain expression archive database. In 2011 IEEE International Conference on Automatic Face & Gesture Recognition (FG) 2011 Mar 21 (pp. 57–64). IEEE. doi: 10.1109/FG.2011.5771462.

32. Kasaeyan Naeini E, Subramanian A, Calderon MD, Zheng K, Dutt N, Liljeberg P, Salantera S, Nelson AM, Rahmani AM. Pain recognition with electrocardiographic features in postoperative patients: method validation study. Journal of Medical Internet Research. 2021 May 28;23(5):e25079. doi:10.2196/25079.

33. Aqajari SA, Cao R, Kasaeyan Naeini E, Calderon MD, Zheng K, Dutt N, Liljeberg P, Salanterä S, Nelson AM, Rahmani AM. Pain assessment tool with electrodermal activity for postoperative patients: method validation study. JMIR mHealth and uHealth. 2021 May 5;9(5):e25258.

34. Cao R, Aqajari SA, Naeini EK, Rahmani AM. Objective pain assessment using wrist-based ppg signals: A respiratory rate based method. In 2021 43rd Annual International Conference of the IEEE Engineering in Medicine & Biology Society (EMBC) 2021 Nov 1 (pp. 1164–1167). IEEE. doi:10.1109/EMBC46164.2021.9630002.

35. Sarker VK, Jiang M, Gia TN, Anzanpour A, Rahmani AM, Liljeberg P. Portable multipurpose bio-signal acquisition and wireless streaming device for wearables. In 2017 IEEE sensors applications symposium (SAS) 2017 Mar 13 (pp. 1–6). IEEE. doi: 10.1109/SAS.2017.7894053.

36. Empatica, “E4 wristband user’s manual,” 2015.

37. Naeini EK, Jiang M, Syrjälä E, Calderon MD, Mieronkoski R, Zheng K, Dutt N, Liljeberg P, Salanterä S, Nelson AM, Rahmani AM. Prospective study evaluating a pain assessment tool in a postoperative environment: protocol for algorithm testing and enhancement. JMIR Research Protocols. 2020 Jul 1;9(7):e17783. doi:10.2196/17783.

38. Gomes PM, Margaritoff P, Silva H. pyHRV: Development and evaluation of an open-source python toolbox for heart rate variability (HRV). In Proc. Int’l conf. On electrical, electronic and computing engineering (icetran) 2019 Jun (pp. 822–828).

39. Laitala J, Jiang M, Syrjälä E, Naeini EK, Airola A, Rahmani AM, Dutt ND, Liljeberg P. Robust ECG R-peak detection using LSTM. In Proceedings of the 35th annual ACM symposium on applied computing 2020 Mar 30 (pp. 1104–1111).

40. Aqajari SA, Naeini EK, Mehrabadi MA, Labbaf S, Dutt N, Rahmani AM. pyeda: An open-source python toolkit for pre-processing and feature extraction of electrodermal activity. Procedia Computer Science. 2021 Jan 1;184:99–106.

41. Greco A, Valenza G, Lanata A, Scilingo EP, Citi L. cvxEDA: A convex optimization approach to electrodermal activity processing. IEEE Transactions on Biomedical Engineering. 2015 Aug 28;63(4):797–804. doi: 10.1109/TBME.2015.2474131.

42. Madhav KV, Ram MR, Krishna EH, Komalla NR, Reddy KA. Estimation of respiration rate from ECG, BP and PPG signals using empirical mode decomposition. In 2011 IEEE International instrumentation and measurement technology conference 2011 May 10 (pp. 1–4). IEEE. doi: 10.1109/IMTC.2011.5944249.

43. Schmidhuber J. Deep learning in neural networks: An overview. Neural networks. 2015 Jan 1;61:85–117.

44. Le QV. A tutorial on deep learning part 2: Autoencoders, convolutional neural networks and recurrent neural networks. Google Brain. 2015 Oct 20;20:1–20.

45. Zhang Z. Improved adam optimizer for deep neural networks. In 2018 IEEE/ACM 26th international symposium on quality of service (IWQoS) 2018 Jun 4 (pp. 1–2). Ieee. doi: 10.1109/IWQoS.2018.8624183.

46. Chawla NV, Bowyer KW, Hall LO, Kegelmeyer WP. SMOTE: synthetic minority over-sampling technique. Journal of artificial intelligence research. 2002 Jun 1;16:321–57.

47. Lemaître G, Nogueira F, Aridas CK. Imbalanced-learn: A python toolbox to tackle the curse of imbalanced datasets in machine learning. The Journal of Machine Learning Research. 2017 Jan 1;18(1):559–63.

48. Ratner A, Bach SH, Ehrenberg H, Fries J, Wu S, Ré C. Snorkel: Rapid training data creation with weak supervision. In Proceedings of the VLDB Endowment. International Conference on Very Large Data Bases 2017 Nov (Vol. 11, No. 3, p. 269). NIH Public Access. doi: 10.1007/s00778-019-00552-1.

49. Kohavi R. A study of cross-validation and bootstrap for accuracy estimation and model selection. In Ijcai 1995 Aug 20 (Vol. 14, No. 2, pp. 1137–1145).

50. Gunes H, Piccardi M. Affect recognition from face and body: early fusion vs. late fusion. In 2005 IEEE international conference on systems, man and cybernetics 2005 Oct 12 (Vol. 4, pp. 3437–3443). IEEE. doi: 10.1109/ICSMC.2005.1571679.

51. A. Ross, Fusion, Feature-Level, pp. 597–602. Boston, MA: Springer US, 2009.

52. Naeini EK, Shahhosseini S, Kanduri A, Liljeberg P, Rahmani AM, Dutt N. AMSER: Adaptive Multimodal Sensing for Energy Efficient and Resilient eHealth Systems. In 2022 Design, Automation & Test in Europe Conference & Exhibition (DATE) 2022 Mar 14 (pp. 1455–1460). IEEE.

53. Taylor S, Jaques N, Nosakhare E, Sano A, Picard R. Personalized multitask learning for predicting tomorrow’s mood, stress, and health. IEEE Transactions on Affective Computing. 2017 Dec 19;11(2):200–13. doi: 10.1109/TAFFC.2017.2784832.

